# Quantifying behavior change during the first year of the COVID-19 pandemic in the United States

**DOI:** 10.1101/2022.01.10.22268799

**Authors:** Dennis L Chao, Victor Cho, Amanda S Izzo, Joshua L Proctor, Marita Zimmermann

## Abstract

**Background:** During the first year of the COVID-19 pandemic, the most effective way to reduce transmission and to protect oneself was to reduce contact with others. However, it is unclear how behavior changed, despite numerous surveys about peoples’ attitudes and actions during the pandemic and public health efforts to influence behavior.

**Methods:** We used two sources of data to quantify changes in behavior at the county level during the first year of the pandemic in the United States: aggregated mobile device (smartphone) location data to approximate the fraction of people staying at home each day and digital invitation data to capture the number and size of social gatherings.

**Results:** Between mid-March to early April 2020, the number of events fell and the fraction of devices staying at home peaked, independently of when states issued emergency orders or stay-at-home recommendations. Activity began to recover in May or June, with later rebounds in counties that suffered an early spring wave of reported COVID-19 cases. Counties with high incidence in the summer had more events, higher mobility, and less stringent state-level COVID-related restrictions the month before than counties with low incidence. Counties with high incidence in early fall stayed at home less and had less stringent state-level COVID-related restrictions in October, when cases began to rise in some parts of the US. During the early months of the pandemic, the number of events was inversely correlated with the fraction of devices staying at home, but after the fall of 2020 mobility appeared to stay constant as the number of events fell. Greater changes in behavior were observed in counties where a larger fraction voted for Biden in the 2020 US Presidential election. The number of people invited per event dropped gradually throughout the first year of the pandemic.

**Conclusions:** The mobility and events datasets uncovered different kinds of behavioral responses to the pandemic. Our results indicate that people did in fact change their behavior in ways that likely reduced COVID exposure and transmission, though the degree of change appeared to be affected by political views. Though the mobility data captured the initial massive behavior changes in the first months of the pandemic, the digital invitation data, presented here for the first time, continued to show large changes in behavior later in the first year of the pandemic.

## Introduction

During the first year of the COVID-19 pandemic in the United States, social distancing was the most effective way to reduce transmission and to protect oneself [Grantz et al 2020]. Many policies were enacted to encourage people to stay home and to limit the size of gatherings [Moreland et al 2020, Hallas et al 2021], though other factors such as rising COVID cases and public messaging also caused people to avoid contact with others [Buckee et al 2021]. Adherence to social distancing took a toll on society, and adherence to safer behaviors seemed to wane over time [Pan et al 2020, Petherick et al 2021].

Social distancing likely reduced transmission of SARS-CoV-2 but is difficult to measure. Self-reported behavior during the pandemic is expensive to collect and might not be reliable. Conversely, mobile device (i.e., smartphone) location data can be used to quantitatively capture the movement of people [Kishore et al 2020, Persson et al 2021], but they are not well-suited to capturing small social gatherings, which may play a large role in the spread of SARS-CoV-2 [Tupper et al 2020, Whaley et al 2021]. However, digital invitation services can be used to collect data on the size and frequency of social gatherings. By combining epidemiologic data with both mobility and digital invitation data, we may begin to quantify the relationships between COVID-19 cases, policies, and individual behavior.

We hypothesize that behavior change can be measured using mobility and digital invitations, and behavior changes are driven by COVID-19 case trends and policies. Here, we use aggregated mobile device data from SafeGraph to quantify population mobility [SafeGraph 2020] and digital invitation data from Evite to capture trends in social gatherings. We also analyze how local factors, such as median income and voting in the 2020 presidential election, correlate with behavior change in response to the pandemic. We aim to quantify behavior changes across the country during the first year of the pandemic in order to inform policymakers for future public health crises.

## Data and methods

### Data

The numbers of reported COVID cases and deaths at the county level were obtained from Johns Hopkin’s Coronavirus Research Center from January 2019 through March 2021 [Johns Hopkins University 2021]. The numbers are reported cumulatively, and our estimates of daily cases and deaths were computed by subtracting the previous day’s cumulative estimate from the current day’s.

We assessed the number and size of events using event data from Evite from Jan 1, 2019 to March 11, 2021. Event data included the date the event was planned (created online), the date of the event, the event type (category), the number of guests invited, the number who RSVPed, and the event location (zip code and/or state). This dataset does not contain information about the individuals who organized or were invited to events. We defined “held” events to be events that were not “virtual” and that were not canceled. When possible, we assigned each event to a county by identifying the county that had the most overlap with the event zip code, using shapefiles for US counties and ZCTAs (https://www.census.gov/geographies/mapping-files/time-series/geo/cartographic-boundary.html). Events that could not be linked to a state (or the District of Columbia) were removed. We defined two time windows for some analyses: “2019” (all events that were to take place in 2019) and “pandemic” (events from March 12 2020-March 11 2021). We restricted county-level analyses to counties with at least 500 Evite events in 2019.

SafeGraph produces aggregated, anonymized datasets based on mobile device location data. We obtained estimates of the number of cell phones based in each census block group in the United States and the number of them that were not detected leaving their imputed “home” location each day from January 2019 through March 2021, which allows us to estimate the fraction of people who stay at home each day [SafeGraph 2020]. Data were obtained by joining SafeGraph’s COVID-19 Data Consortium (https://www.safegraph.com/covid-19-data-consortium).

Population and median income estimates for US counties in 2019 were obtained from the American Community Survey product of the United States Census, accessed using the R package tidycensus [Walker et al 2020]. We used the 2013 NCHS Urban-Rural Classification Scheme for Counties (https://www.cdc.gov/nchs/data_access/urban_rural.htm) to identify urban, suburban, and rural counties.

The dates of state-level emergency and stay-at-home orders in response to COVID-19 were obtained from CUSP, available at https://statepolicies.com.

Data on state-level policy “stringency” and gathering policies related to COVID-19 were obtained from the Oxford COVID-19 Government Response Tracker [Hale et al 2021, Hallas et al 2021], which is available at https://www.bsg.ox.ac.uk/news/our-coronavirus-policy-tracker-adds-data-us-states.

Data on 2020 US election results was obtained from the MIT Voting data from the Massachusetts Institute of Technology Election Data + Science Lab (https://electionlab.mit.edu/data). The county-level 2020 presidential election results are available at https://doi.org/10.7910/DVN/VOQCHQ.

## Methods

We analyzed aggregated data from mobile device locations and from a digital invitation service in order to provide complementary views of human behavior. We first tabulated events scheduled, held, and canceled in 2019 and pandemic by county per capita, day of the week, holiday, event type, and event size. For event size, we examined the number of people invited as well as the fraction who RSVPed “yes.” Finally, we assess the planning window, or time between sending the invite and the event date. Similarly, we tabulated the number of cell phones staying completely at home by day of the week.

We defined three “waves” of reported COVID-19 cases: a spring wave in March-April 2020, a summer wave in June-August 2020, and an early fall wave in October 2020. We then categorized counties as experiencing each of these waves or not. Counties with a spring or summer wave were the 30 counties with the highest number of reported COVID-19 cases per capita during each of these two-month time periods, respectively, whereas counties with no wave were the 200 counties with the lowest incidence during the time period. Because the fall wave was experienced nationally, we defined early vs late fall wave as having the highest vs lowest incidence in October 2020. Over the same time periods, we aggregated when gathering size policies, as defined as “stringency” above, were implemented, loosened, or strengthened.

We compared measures of behavior and policy stringency between counties with high and low reported COVID-19 case incidence using the Wilcoxon rank-sum test. The Wilcoxon rank-sum test was used to compare the distributions of the numbers of invited guests to events before and after gathering restrictions were declared.

We used linear regression to quantify the relationship between votes in the 2020 US Presidential election and behavior. We fit models to predict the county-level behavioral metric (drop in events or change in mobility) using the fraction of votes for Biden in the 2020 US Presidential election. To determine if the state governor’s affiliation played a significant role in this relationship, we used stepwise Akaike’s Information Criterion (AIC) to determine if affiliation had a significant effect on the intercept or slope of the model. To determine if different states had different relationships between election results and behavior, we used AIC to determine if the state (as a categorical variable) should be included in the model.

Analyses were performed using R version 4.0.5 [R Core Team 2021].

## Results

### Trends in COVID cases and policies

COVID cases increased throughout the epidemic over the course of the three waves (Figure 1A). Some regions had waves of COVID-19 cases in the spring of 2020, others in the summer, and the entire US experienced a wave starting in the fall or early winter.

**Figure 1.**
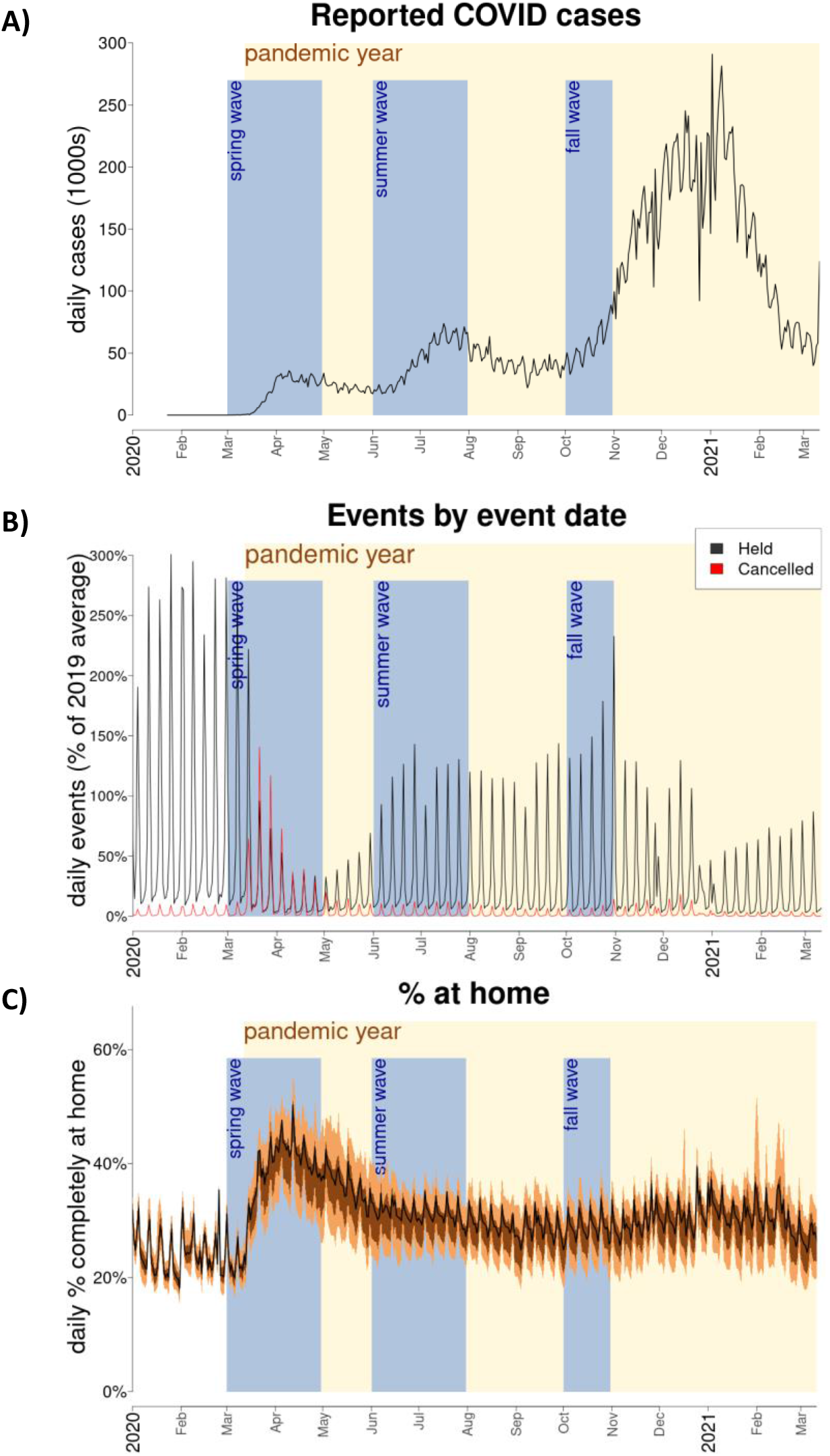
COVID-19 cases, events, and mobility. A) Daily number of reported COVID-19 cases in the United States. B) Daily number of Evite events by event date. The black line represents the daily number of events held as a percentage of the average number of events scheduled for each day in 2019. The red line represents the number of events that were canceled. C) Percent of mobile devices home each day. The light brown region covers 90% of the daily range of all counties, and the dark brown covers 50% of the range. The solid line is the average value for the United States. In all panels, time windows used in the analyses are highlighted: the pandemic year (March 12, 2020-March 11, 2021, highlighted in yellow), the spring wave (March-April 2020, in blue), the summer wave (June-July 2020, in blue), and the start of the fall wave (October 2020, in blue).

Most states (and some counties) had guidance on gathering size that changed over time. In March 2020, 42 states (and the District of Columbia) enacted gathering size restrictions (Figure S1). Between late April and early July, most states (31) loosened restrictions. In mid-July to early August, 6 states tightened restrictions again. In November - December, many states enacted restrictions. Several states loosened restrictions in February 2021.

### Trends in events

In total, we included 5,400,380 Evite events from the 50 states or the District of Columbia in 2019 and 1,788,709 during the “pandemic period,” and 5,245,710 in 2019 and 1,727,546 linked with zip codes in a total of 571 counties (Table 1). Most Evite events were from urban (>2 million events in 2019, or 44.7% of events) and suburban counties (nearly 2 million events in 2019, or 37.5% of events), and the highest number of events per capita were from suburban counties (24.1 events per 1000 pop per year in 2019) followed by urban (23.2 events per 1000).

**Table 1.** Evite events by urbanization classification by county. “Pandemic” period covers events scheduled from March 12 2020-March 11 2021, while “2019” is from Jan 1 2019-Dec 31 2019.

|  | Evite events N in millions (%) |  | Events per 1000 pop |  |
| --- | --- | --- | --- | --- |
|  | 2019 | pandemic | 2019 | pandemic |
| **Urbanization** |  |  |  |  |
| Large central metro | 2.32M (44.7%) | 0.70M (41.0%) | 23.2 | 7.0 |
| Large fringe metro<br>(suburban) | 1.95M (37.5%) | 0.65M (38.5%) | 24.1 | 8.1 |
| Medium metro | 0.66M (12.7%) | 0.24M (14.0%) | 9.7 | 1.4 |
| Small metro | 0.15M (3.0%) | 0.06M (3.5%) | 5.2 | 0.8 |
| Micropolitan | 0.08M (1.5%) | 0.03M (2.0%) | 2.8 | 0.4 |
| Noncore (rural) | 0.03M (0.6%) | 0.02M (1.0%) | 1.7 | 0.2 |
| **Day of week** |  |  |  |  |
| Saturdays | 2.58M (47.8%) | 0.92M (51.7%) | 8.0 | 2.8 |
| Sundays | 1.20M (22.2%) | 0.21M (19.4%) | 3.7 | 1.1 |
| weekdays | 1.62M (30.0%) | 0.52M (29.0%) | 5.0 | 1.6 |

Most Evite events were scheduled to take place on weekends, with 47.8% of events on Saturdays, 22.2% on Sundays, and 12.1% on Fridays in 2019 (Table 1). Events occurred throughout the year, but there were conspicuous spikes around major holidays, such as Christmas and Halloween (Figure 1B, Figure S2). There were 135 categories of events, with “Birthday for Kids” being the most popular event type (1,190,545 events in 2019, or 22% of all events).

Across the country, fewer events were planned after the 2^nd^ week of March 2020 (Figure S3). Many events scheduled to occur during the 2^nd^ - 3^rd^ weeks of March were cancelled (Figure 1B). The combination of reduced event scheduling and widespread event cancellation resulted in a rapid drop in events held during the 3^rd^ weekend in March (Figure 1B). The number of events scheduled began dropping before these restrictions were enacted and continued to drop (Figure S3).

The average number of guests invited to Evite events during the first year of the pandemic was lower than in 2019 across most categories of events (Figure S4, Figure S5A), leading to a lower number RSVPing “yes” per event (Figure S5B). However, the fraction of invitees RSVPing “yes” was similar during 2019 and 2020 (Figure S5C).

Average event size decreased over the course of the pandemic. Event categories that typically had large numbers of invited guests in 2019 (fundraisers, open houses, professional events) dropped more than typically smaller categories (Figure S6). The planning window between event creation and event date also decreased. Evites had typically been created 18 days before events, but this planning window dropped to 14 days during the pandemic (Figure S7).

### Trends in mobility

Before the pandemic, the fraction of mobile devices that stayed at home each day was lowest on weekdays (22.8%) and highest on weekends (26.5% on Saturdays and 29.9% on Sundays) (Table 2). The fraction of people staying at home each day was highest in March-April 2020 and returned to near pre-pandemic levels on weekends but remained relatively high on weekdays by the fall of 2020 (Figure 1C, Table 2).

**Table 2.** Mobility by urbanization classification by county and date. The numbers represent the average % of mobile devices staying at completely at home each day during the time periods specified by the columns.

|  | Average % staying at home |  |  |  |
| --- | --- | --- | --- | --- |
|  | Jan-Feb 2020 | Mar-Apr 2020 | Jun-Jul 2020 | Oct 2020 |
| **Urbanization** |  |  |  |  |
| Large central metro | 25.1 | 39.7 | 35.1 | 31.4 |
| Large fringe metro<br>(suburban) | 23.3 | 38.6 | 31.6 | 29.2 |
| Medium metro | 24.4 | 34.9 | 30.1 | 27.8 |
| Small metro | 24.4 | 32.8 | 28.1 | 26.2 |
| Micropolitan | 24.1 | 31.0 | 26.7 | 25.0 |
| Noncore (rural) | 24.1 | 29.3 | 25.7 | 24.0 |
| **Day of week** |  |  |  |  |
| Saturdays | 26.5 | 38.1 | 30.8 | 27.5 |
| Sundays | 29.9 | 40.5 | 34.5 | 32.7 |
| weekdays | 22.8 | 35.4 | 30.8 | 28.2 |

### Correlations between number of events and mobility

Both before and during the pandemic, Evite events were held mostly on weekends while mobile devices stayed completely at home the most on weekends (Tables 1 and 2). As cases were reported in the United States in March 2020, the number of events dropped and the fraction of mobile devices staying at home rose (Figures 1B-C). The number of events reached a low and the fraction staying at home reached a high in April 2020 (Figure 3). Both metrics partially returned to pre-pandemic levels in the summer of 2020, then the fraction staying at home stayed relatively constant while the number of events briefly spiked in late October 2020, reached new lows in January 2021 (Figure 3). On some dates, like Superbowl Sunday on February 2, 2020, and Halloween on the last weekend of October 2020, the number of events spikes without obvious changes in the fraction staying at home (Figure S2).

**Figure 3.**
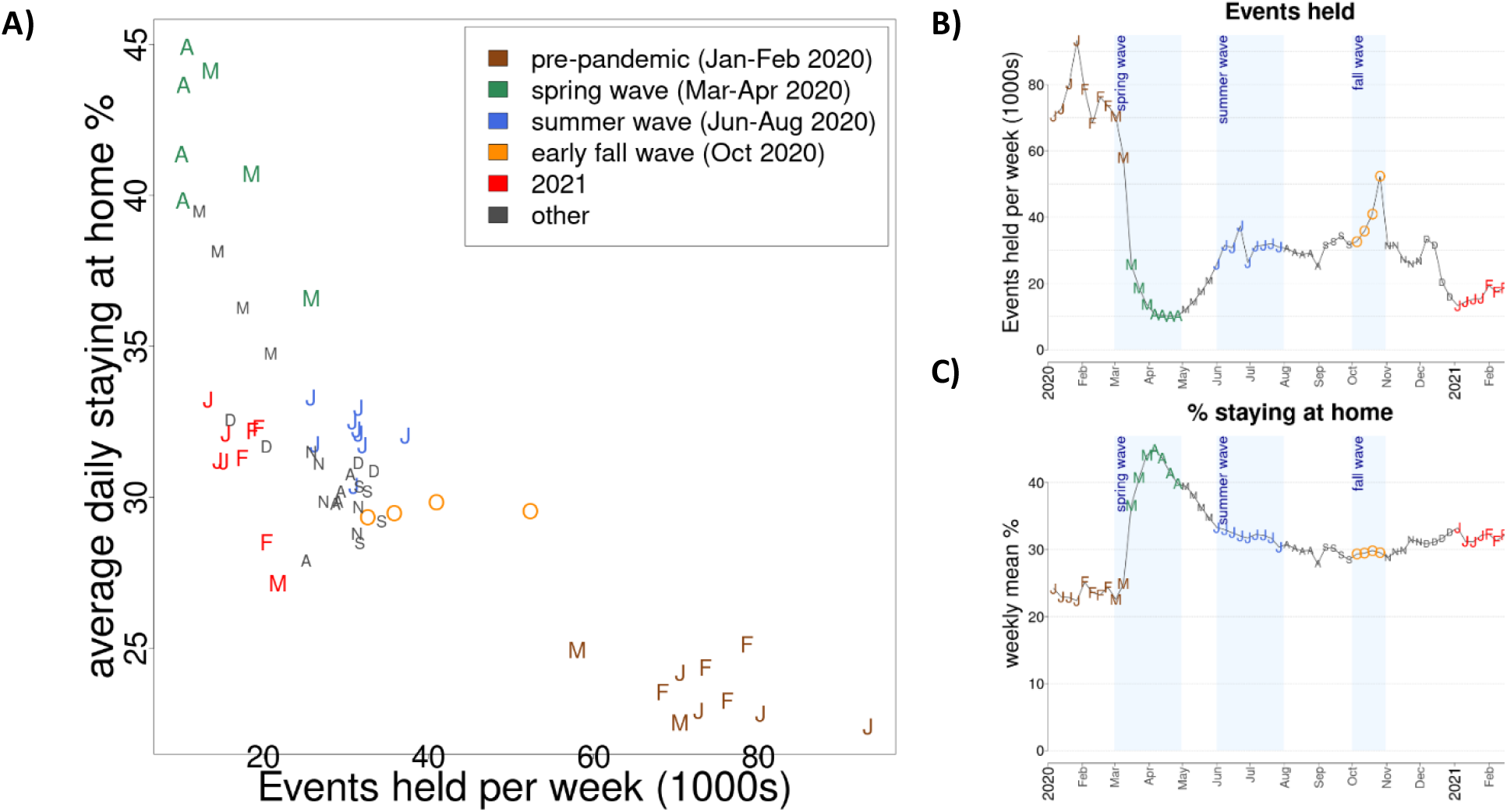
Number of events vs % staying at home each week. A) The scatterplot represents weekly numbers of events on the x-axis and the average fraction of mobile devices staying at home each day during corresponding weeks. Each point is plotted as the first letter of the month that begins each week (i.e., “J” for January, June, and July). Colors indicate different phases of the pandemic, as indicated in the legend. B) The weekly number of Evite events held. C) The weekly average % of mobile devices that stay completely at home each day.

### Comparison between COVID-19 incidence and behavior

In counties with spring waves of reported COVID-19 cases, Evite activity remained low throughout April and May before climbing in June. A high percentage of people in these counties stayed completely at home in April (Figure 4, Table S3). Policy stringency remained high in the states with spring waves. In counties with low spring incidence, the number of Evite events began to climb earlier (in late May) and the fraction of people who stayed home was lower compared to counties with high spring waves (p<0.001, Wilcoxon rank-sum test). Counties with summer waves (defined as high incidence in June and July) had more events (p<0.001), stayed home less (p<0.001), and had less stringent policies (p<0.001) before the summer peak (May-June 2020) compared to counties with low summer incidence. Counties with high incidence in October (i.e., where the fall wave began early) stayed at home less and had less stringent state policies in October than counties where the fall wave struck later.

**Figure 4.**
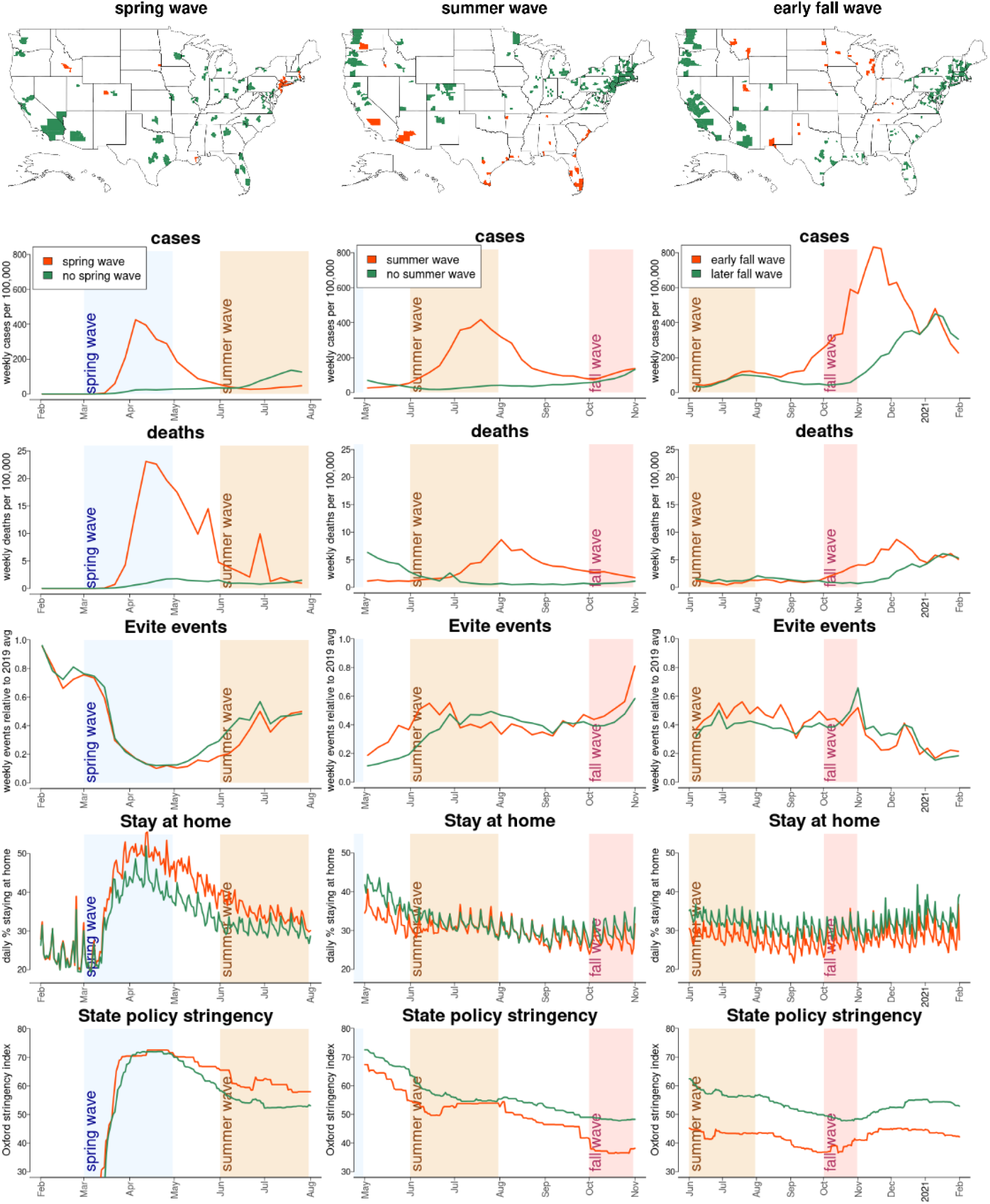
Characteristics of counties with high COVID-19 incidence during spring, summer, or early fall. The top row shows which counties had spring waves of reported COVID-19 cases (left), summer waves (middle), and early fall waves (right) in orange. Counties in green had low per capita incidence during these time periods. The panels below the maps plot the number of cases per capita during the relevant time period, the COVID deaths per capita, relative number of events, the fraction of mobile devices staying completely at home, and the stringency of state policies. Lines in orange are for counties with high incidence and green are for low incidence (matching the map colors).

We identified six states where all gatherings (i.e., indoors and outdoors) were restricted to 10 or fewer people between July and mid-December and where there were enough events for analysis (which excludes medium and small states) (Table S4). These restrictions should have impacted Evite events, which often have more than 10 invited guests (Figures 5 and S8). The number of guests invited per event was significantly lower in some of these states in the two weeks after restrictions compared to the two weeks before (Figure 5). However, in some states the number of events appeared to decline before restrictions (Figure S8).

**Figure 5.**
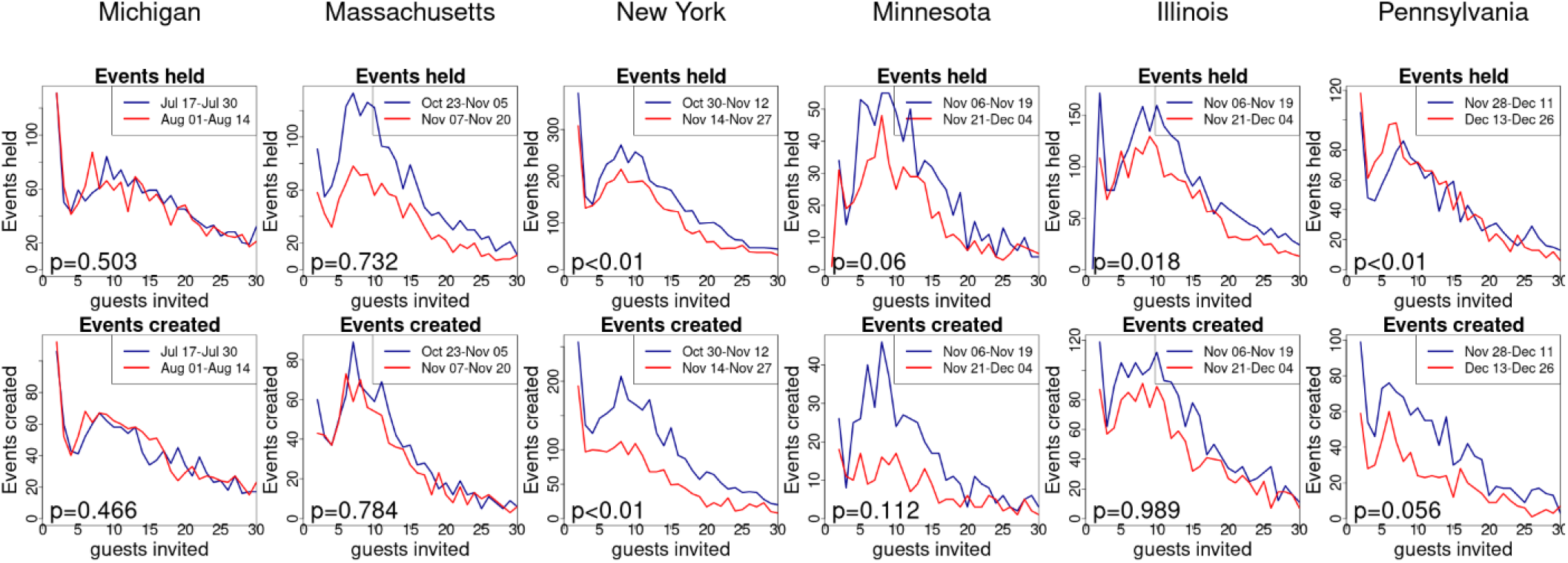
Response to tightening gathering restrictions. Each column represents a state that restricted gatherings to 10 people or fewer than 10 people. Each line plots the number of events (y-axis) by the number of guests invited to the events (x-axis). The blue lines are for events that were held (top panels) or created (bottom panels) in the two weeks before gathering restrictions, while the red lines are for events in the two weeks after. The p-values from comparing the distributions of the number of guests in the two weeks before and the two weeks after restrictions are printed in the lower left of each panel.

### County-level predictors of behavior change

We analyzed the relationship between political views (using voting for Biden in the 2020 US Presidential election as a proxy) and behavior changes in response to the pandemic. The number of events dropped more during the first pandemic year (March 12, 2020 – March 11, 2021) in counties with a higher proportion of votes for Biden in the 2020 presidential election (slope=-0.48, R^2=0.35, Figure 6A). There were no significant differences in slope among states but there were differences in intercept (Figure S9). States with Republican governors had a small but statistically significant 3.8% smaller drop in events during the pandemic compared to Democratic-led states (p<0.0001). The fraction of devices staying completely at home rose in March and April of 2020 then fell in the summer, but not to pre-pandemic levels. The fraction of devices staying at home was correlated with the proportion voting for Biden both before and during the pandemic, but the difference between counties supporting Biden and those that did not grew larger between pre-pandemic February 2020 and July of 2020 (Figure S10; for every additional 1% of voters supporting Biden, an additional 0.056% stayed at home in February and 0.176% in July). The difference in fraction staying home between July and February was also positively correlated with the fraction voting for Biden (Figures 6B, S11).

**Figure 6.**
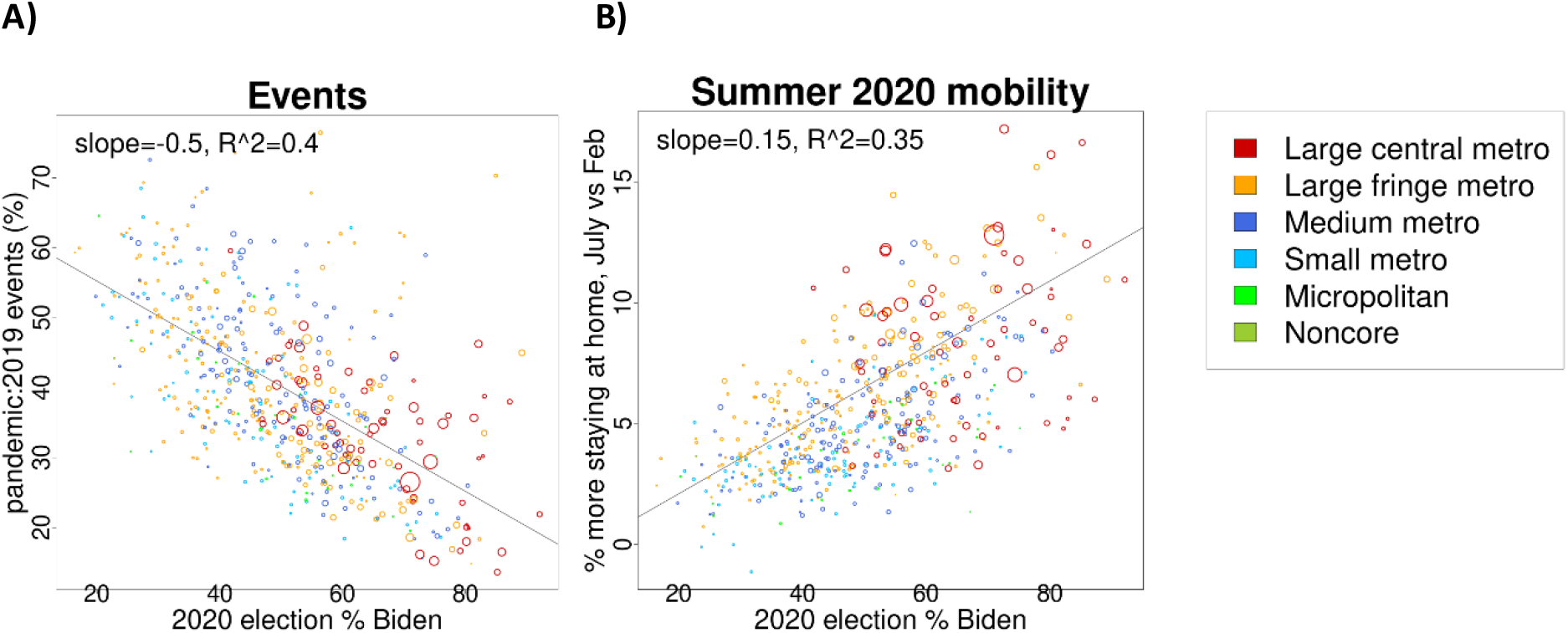
Drop in events and increase in staying at home vs fraction voting for Biden in the 2020 election by county. A) Number of events vs the fraction voting for Biden. The y-axis plots the ratio of events during the pandemic (March 2020-2021) vs 2019. Dot sizes are proportional to county population and colored according to the urban/rural classification indicated in the legend. Only counties with at least 500 Evites in 2019 are plotted. The solid gray line is the linear regression fit. B) Mobility in July 2020 vs the fraction voting for Biden. The y-axis plots the average daily fraction of mobile devices staying home in July 2020 minus the average in February 2020.

Higher-income counties had more Evites per capita than lower-income in 2019, but the relative decline in Evite events during the pandemic year was larger in counties with higher income both among major metropolitan areas and suburbs (Table S1).

## Discussion

Using Evite event and SafeGraph mobile device location data, we found evidence that people in the United States engaged in social distancing behavior that likely reduced the spread of SARS-CoV-2. During the pandemic, COVID-19 cases, social events, human mobility, policies, and public perception interacted in ways that made it difficult to tease apart specific temporality and hence causality, but we are able to report on several interactions from this unique combination of data sources.

Evite events and SafeGraph aggregated mobile device location data capture different kinds of human activity. Evite activity represents social gatherings, while cell phone mobility might be dominated by going to work and other activities. More mobile devices (and presumably people) stay at home on weekends than weekdays, likely because adults often go to work on weekdays. In contrast, Evite events are social activities that occur mostly on weekends. Therefore, the number of Evite events is highest on days when most people stay at home the most, which includes some holidays. During a public health emergency, increased staying at home is not an indication of increased leisure time associated with social gatherings. The event data provides a novel and quantitative dimension to understanding population interactions that is complementary to cellphone mobility indicators.

In March of 2020, the number of Evite events plummeted at about the same time across the US, in some states before and in others after state-level emergency orders and often well before stay-at-home orders. Badr et al [2020] also found that mobility dropped at about the same time across the US, often before local- or state-level directives. Many people stayed home starting in mid-March. Early in the pandemic, it was likely that high county death rates were associated with the perception of higher risk and more staying at home [Elharake et al 2021]. As COVID case fatality rates declined and “pandemic fatigue” set in, the associations between cases, behavior, and policy become harder to measure. Though the number of events and mobility both dropped early in the pandemic and began to recover in the late spring, mobility plateaued starting in the summer of 2020 while the number of events dropped in the winter. The divergence of trends from these two data sources indicates that they measure different kinds of behavior. Evite events are dominated by small social events, especially weekend birthday parties, while SafeGraph mobility data tracks mobile device locations, which might be dominated by weekday commutes to work.

Government policies have been linked to reductions in COVID-19 cases in the literature [Li et al 2021, Persson et al 2021, Liu et al 2021]. In our study, we observed trends consistent with the hypothesis that rising COVID-19 cases caused people to socially distance and for governments to issue policies to slow transmission. This makes it difficult to determine the contribution of these policies to behavior change. Additionally, we did not observe a consistent reduction in event sizes in response to strict gathering restriction policies, though these policies have been associated with reductions in COVID-19 cases [Persson et al 2021, Liu et al 2021, Li et al 2021]. People have behavioral responses to both COVID cases and policy changes, while at the same time, COVID cases are impacted by behavior, and policies respond to changes in COVID cases. These cyclical causalities make it difficult to quantify individual impacts [Buckee et al 2021]. Further, we see different trends in different parts of the country.

We found that behavior change was associated with candidate preference in the 2020 US Presidential election. We found that behavioral responses to the pandemic seemed attenuated among counties with a smaller proportion of votes for Biden in the 2020 US Presidential election, which is consistent with earlier studies on political affiliation and attitudes about the pandemic [Grossman et al 2020, Gratz et al 2021, Neelon et al 2021, Clinton et al 2021] or mobility during the pandemic [Kavanagh et al 2021].

We also found that the number of events dropped more in counties with higher median income. Some have hypothesized that high-income populations social distance more because they are more likely to have the ability to work from home and use delivery services instead of going to stores [e.g., Weill et al 2020, Jay et al 2020]. While it seems likely that necessity partially explains the higher rates of mobility observed among lower-income regions, the event data show differences by income in social gatherings as well.

There were several limitations to our analysis. First, we assumed that Evites events represented in-person events and cancellation and RSVP data was reliable. Second, we are not able to disentangle the impacts of policy and other drivers of behavior change. Similarly, policy can be local, complicated, and heterogenous, making it difficult to compare nationally. Additionally, messaging from government outside of formal policies was not included here but may have influenced behavior [Pink et al 2021]. We acknowledge that factors we did not include in our analyses, such as weather and school closures, could have played a major role in SARS-CoV-2 transmission. Finally, we use ecological comparisons, and cannot compare individual-level behavior using these methods. We know that Evite users, those tracked by SafeGraph [Squire et al 2020, Grantz et al 2020, SafeGraph 2021], and voters [Verba et al 1995, Igielnik et al 2021] do not represent the general population. While Evites are most used in higher-income suburbs and major metropolitan areas, COVID incidence has been highest in rural and lower income counties [Li et al 2021]. However, the millions of events per year facilitated by Evite represent a unique data source for tracking trends in social gatherings.

The first year of the pandemic enabled us to observe and quantify the relevant processes as well as to determine the appropriate geopolitical scales on which they act. Our analysis demonstrates the complexity of forces that drive behavioral responses to a pandemic. Though we identified correlations between rising reported COVID-19 cases and changes in behavior, they could have occurred through any number of ways, including government policies and messaging [Grossman et al 2020, Neelson et al 2021, Pink et al 2021, Li et al 2021, Liu et al 2021], ads and other messages from the media [Breza et al 2021], social media [Cinelli et al 2020], and communication of community COVID-19 death rates [Elharake et al 2021]. Individuals will integrate these messages in the context of their own personal beliefs [Clinton et al 2021, Gratz et al 2021, Kavanagh et al 2021], the experiences of their friends and family [Epstein et al 2008], and “pandemic fatigue” [Pan et al 2020, Petherick et al 2021]. Better understanding of the drivers that influence behavior change will enable more effective messaging and policy decisions.

## Data Availability

Instructions for obtaining the publicly available data used in the present study are documented in the Methods. The data from SafeGraph, Inc, used in the study are no longer supported or available from the source. The data from Evite, Inc, are available upon reasonable request to the authors.

https://www.census.gov/geographies/mapping-files/time-series/geo/cartographic-boundary.html

https://www.cdc.gov/nchs/data_access/urban_rural.htm

https://statepolicies.com/

https://www.bsg.ox.ac.uk/news/our-coronavirus-policy-tracker-adds-data-us-states

https://doi.org/10.7910/DVN/VOQCHQ

https://www.safegraph.com/covid-19-data-consortium

## Acknowledgments

We thank Patrick Codrington and Saurabh Shetty for processing data from Evite and Jeremy Forman and Roy Burstein for productive discussions.

## References

Badr HS, D. H, Marshall M, Dong E, Squire MM, Gardner LM. Association between mobility patterns and COVID-19 transmission in the USA: a mathematical modelling study. Lancet Infect Dis. 2020 Nov;20(11):1247–1254. doi: 10.1016/S1473-3099(20)30553-3.

Breza E, Stanford FC, Alsan M, Alsan B, Banerjee A, Chandrasekhar AG, Eichmeyer S, Glushko T, Goldsmith-Pinkham P, Holland K, Hoppe E, Karnani M, Liegl S, Loisel T, Ogbu-Nwobodo L, Olken BA, Torres C, Vautrey PL, Warner ET, Wootton S, Duflo E. Effects of a large-scale social media advertising campaign on holiday travel and COVID-19 infections: a cluster randomized controlled trial. Nat Med. 2021 Sep;27(9):1622–1628. doi: 10.1038/s41591-021-01487-3.

Buckee C, Noor A, Sattenspiel L. Thinking clearly about social aspects of infectious disease transmission. Nature. 2021 Jul;595(7866):205–213. doi: 10.1038/s41586-021-03694-x.

Cinelli M, Quattrociocchi W, Galeazzi A, Valensise CM, Brugnoli E, Schmidt AL, Zola P, Zollo F, Scala A. The COVID-19 social media infodemic. Sci Rep. 2020 Oct 6;10(1):16598. doi: 10.1038/s41598-020-73510-5.

Clinton J, Cohen J, Lapinski J, Trussler M. Sci Adv. Partisan pandemic: How partisanship and public health concerns affect individuals’ social mobility during COVID-19. Sci Adv 2021 Jan 6;7(2):eabd7204. doi: 10.1126/sciadv.abd7204.

Elharake JA, Shafiq M, McFadden SM, Malik AA, Omer SB. The Association of COVID-19 Risk Perception, County Death Rates, and Voluntary Health Behaviors among U.S. Adult Population. J Infect Dis. 2021 Mar 10:jiab131. doi: 10.1093/infdis/jiab131.

Epstein JM, Parker J, Cummings D, Hammond RA. Coupled contagion dynamics of fear and disease: mathematical and computational explorations. PLoS One. 2008;3(12):e3955. doi: 10.1371/journal.pone.0003955.

Grantz KH, Meredith HR, Cummings DAT, Metcalf CJE, Grenfell BT, Giles JR, Mehta S, Solomon S, Labrique A, Kishore N, Buckee CO, Wesolowski A. The use of mobile phone data to inform analysis of COVID-19 pandemic epidemiology. Nat Commun. 2020 Sep 30;11(1):4961. doi: 10.1038/s41467-020-18190-5.

Gratz KL, Richmond JR, Woods SE, Dixon-Gordon KL, Scamaldo KM, Rose JP, Tull MT. Adherence to Social Distancing Guidelines Throughout the COVID-19 Pandemic: The Roles of Pseudoscientific Beliefs, Trust, Political Party Affiliation, and Risk Perceptions. Ann Behav Med. 2021 May 6;55(5):399–412. doi: 10.1093/abm/kaab024.

Grossman G, Kim S, Rexer JM, Thirumurthy H. Political partisanship influences behavioral responses to governors’ recommendations for COVID-19 prevention in the United States. Proc Natl Acad Sci U S A. 2020 Sep 29;117(39):24144–24153. doi: 10.1073/pnas.2007835117.

Hale T, Angrist N, Goldszmidt R, Kira B, Petherick A, Phillips T, Webster S, Cameron-Blake E, Hallas L, Majumdar S, and Tatlow H. (2021). A global panel database of pandemic policies (Oxford COVID-19 Government Response Tracker). Nat Hum Behav. 2021 Apr;5(4):529–538. doi: 10.1038/s41562-021-01079-8.

Hallas L, Hale T, Hatibie A, Majumdar S, Pyarali M, Koch R, Wood A. Variation in US states’ responses to COVID-19: Blavatnik School working paper. Version 3. Available at: https://www.bsg.ox.ac.uk/research/publications/variation-us-states-responses-covid-19.

Igielnik R, Keeter S, Hartig H. Behind Biden’s 2020 Victory: An examination of the 2020 electorate, based on validated voters. Pew Research Center. 2021. Available at: https://www.pewresearch.org/politics/2021/06/30/behind-bidens-2020-victory/

Johns Hopkins University. COVID-19 Data Repository by the Center for Systems Science and Engineering (CSSE) at Johns Hopkins University. 2021. Available at https://github.com/CSSEGISandData/COVID-19/tree/master/csse_covid_19_data/csse_covid_19_daily_reports.

Kavanagh NM, Goel RR, Venkataramani AS. County-Level Socioeconomic and Political Predictors of Distancing for COVID-19. Am J Prev Med. 2021 Jul;61(1):13–19. doi: 10.1016/j.amepre.2021.01.040.

Kishore N, Kiang MV, Engø-Monsen K, Vembar N, Schroeder A, Balsari S, Buckee CO. Measuring mobility to monitor travel and physical distancing interventions: a common framework for mobile phone data analysis. Lancet Digit Health. 2020 Nov;2(11):e622–e628. doi: 10.1016/S2589-7500(20)30193-X.

Li D, Gaynor SM, Quick C, Chen JT, Stephenson BJK, Coull BA, Lin X. Identifying US county-level characteristics associated with high COVID-19 burden. BMC Public Health. 2021 May 28;21(1):1007. doi: 10.1186/s12889-021-11060-9.

Li Y, Campbell H, Kulkarni D, Harpur A, Nundy M, Wang X, Nair H; Usher Network for COVID-19 Evidence Reviews (UNCOVER) group. The temporal association of introducing and lifting non-pharmaceutical interventions with the time-varying reproduction number (R) of SARS-CoV-2: a modelling study across 131 countries. Lancet Infect Dis. 2021 Feb;21(2):193–202. doi: 10.1016/S1473-3099(20)30785-4.

Liu Y, Morgenstern C, Kelly J, Lowe R; CMMID COVID-19 Working Group, Jit M. The impact of non-pharmaceutical interventions on SARS-CoV-2 transmission across 130 countries and territories. BMC Med. 2021 Feb 5;19(1):40. doi: 10.1186/s12916-020-01872-8.

MIT Election Data and Science Lab. County Presidential Election Returns 2000-2020. Harvard Dataverse, V9. 2021. doi: 10.7910/DVN/VOQCHQ.

Neelon B, Mutiso F, Mueller NT, Pearce JL, Benjamin-Neelon SE. Associations Between Governor Political Affiliation and COVID-19 Cases, Deaths, and Testing in the U.S. Am J Prev Med. 2021 Jul;61(1):115–119. doi: 10.1016/j.amepre.2021.01.034.

Pan Y, Darzi A, Kabiri A, Zhao G, Luo W, Xiong C, Zhang L. Quantifying human mobility behaviour changes during the COVID-19 outbreak in the United States. Sci Rep. 2020 Nov 26;10(1):20742. doi: 10.1038/s41598-020-77751-2.

Persson J, Parie JF, Feuerriegel S. Monitoring the COVID-19 epidemic with nationwide telecommunication data. Proc Natl Acad Sci U S A. 2021 Jun 29;118(26):e2100664118. doi: 10.1073/pnas.2100664118.

Petherick A, Goldszmidt R, Andrade EB, Furst R, Hale T, Pott A, Wood A. A worldwide assessment of changes in adherence to COVID-19 protective behaviours and hypothesized pandemic fatigue. Nat Hum Behav. 2021 Sep;5(9):1145–1160. doi: 10.1038/s41562-021-01181-x.

Pink SL, Chu J, Druckman JN, Rand DG, Willer R. Elite party cues increase vaccination intentions among Republicans. Proc Natl Acad Sci U S A. 2021 Aug 10;118(32):e2106559118. doi: 10.1073/pnas.2106559118.

R Core Team. R: A Language and Environment for Statistical Computing. R Foundation for Statistical Computing. Vienna, Austria. 2021. Available at https://www.R-project.org.

SafeGraph. Data analysis methodology for the SafeGraph stay-at-home index. 2020. https://docs.google.com/document/d/1k_9LGQn95P5gHsSeuBdzgtEWGGCmzXdcOkcphWi0Cas/edit?usp=sharing.

SafeGraph. Privacy policy. 2021. Available at https://www.safegraph.com/privacy-policy. Accessed on May 18, 2021.

Squire, RF. What about bias in the SafeGraph dataset? SafeGraph. 2019. Available at https://www.safegraph.com/blog/what-about-bias-in-the-safegraph-dataset

Tupper P, Boury H, Yerlanov M, Colijn C. Event-specific interventions to minimize COVID-19 transmission. Proc Natl Acad Sci U S A. 2020 Dec 15;117(50):32038–32045. doi: 10.1073/pnas.2019324117.

Verba S, Schlozman KL, and Brady HE. Voice and equality: Civic voluntarism in American politics, Harvard University Press. 1995.

Walker, K. tidycensus: Load US Census Boundary and Attribute Data as ‘tidyverse’ and ‘sf’-Ready Data Frames, R package version 0.9.9.2 (CRAN, 2020); https://CRAN.R-project.org/package=tidycensus

Weill JA, Stigler M, Deschenes O, Springborn MR. Social distancing responses to COVID-19 emergency declarations strongly differentiated by income. Proc Natl Acad Sci U S A. 2020 Aug 18;117(33):19658–19660. doi: 10.1073/pnas.2009412117.

Whaley CM, Cantor J, Pera M, Jena AB. Assessing the Association Between Social Gatherings and COVID-19 Risk Using Birthdays. JAMA Intern Med. 2021 Aug 1;181(8):1090–1099. doi: 10.1001/jamainternmed.2021.2915.

