## Supplemental Tables and Figures for "Quantifying behavior change during the first year of the COVID-19 pandemic in the United States"

### Supplemental figures and tables

**Table S1. Evite events by median income and urban classification by county.** Income quartiles were defined within urbanization categories and are population-weighted.

| | Evite events 2019 (in millions) | | 2019 Events per 1000 pop | | Ratio of pandemic:2019 events | |
|---|---|---|---|---|---|---|
| Median income | Large central metro | Large fringe metro | Large central metro | Large fringe metro | Large central metro | Large fringe metro |
| 1st quartile (lowest) | 0.35M | 0.20M | 13.9 | 10.0 | 0.387 | 0.456 |
| 2nd quartile | 0.56M | 0.38M | 20.4 | 18.9 | 0.338 | 0.378 |
| 3rd quartile | 0.53M | 0.58M | 23.1 | 28.0 | 0.304 | 0.344 |
| 4th quartile | 0.89M | 0.79M | 36.1 | 39.4 | 0.245 | 0.282 |

**Table S2. Number of counties by urbanization and the fraction with >=500 Evite events in 2019.**

| ***urbanization*** | ***Counties with >=500 events in 2019*** | ***Total counties in US*** | ***% with >=500 events*** |
|---|---|---|---|
| Large central metro | 68 | 68 | 100% |
| Large fringe metro (suburb) | 217 | 368 | 59.0% |
| Medium metro | 166 | 373 | 44.5% |
| Small metro | 90 | 358 | 25.1% |
| Micropolitan | 29 | 641 | 4.5% |
| Noncore (rural) | 1 | 1341 | 0.1% |

**Table S3. Comparison of counties with high spring, summer, or early fall incidence.** Numbers indicate the mean and range of values at the county level. p-values from Wilcoxon rank sum test. A few counties have negative numbers of cases and deaths during some time periods due to reporting artifacts.

| | March-April 2020 | | | June-July 2020 | | | October 2020 | | |
|---|---|---|---|---|---|---|---|---|---|
| | Spring peak (N=30) | No spring peak (N=200) | p | Summer peak (N=30) | No summer peak (N=200) | p | Early fall wave (N=30) | Later fall wave (N=200) | p |
| cases/1000 | 18.0 (10.0-36.1) | 1.3 (0.2-2.9) | - | 23.6 (18.6-46.7) | 2.4 (0.0-4.0) | - | 19.3 (13.0-33.3) | 2.4 (0.0-3.5) | - |
| deaths/1000 | 0.96 (0.04-2.16) | 0.06 (0.00-0.39) | - | 0.31 (0.11-1.14) | 0.11 (-0.14-0.62) | - | 0.14 (0.03-0.30) | 0.04 (0.00-0.35) | - |
| Evites compared to 2019 avg | 0.30 (0.16-0.38) | 0.31 (0.19-0.51) | 0.47 | 0.43 (0.23-1.03) | 0.39 (0.07-0.85) | 0.28 | 0.49 (0.21-0.96) | 0.51 (0.13-1.09) | 0.54 |
| Mean stay-at-home % | 41.8 (32.9-49.7) | 36.5 (28.8-45.8) | <0.001 | 30.8 (23.8-35.8) | 31.5 (22.8-42.9) | 0.45 | 27.6 (21.4-32.7) | 29.7 (18.0-37.2) | <0.001 |
| Mean state stringency | 54.8 (44.7-63.9) | 52.4 (38.6-63.9) | 0.21 | 52.9 (37.1-63.8) | 56.7 (30.7-80.6) | 0.15 | 38.5 (11.8-71.7) | 48.5 (25.6-69.8) | <0.001 |

**Table S4. Selected state-issued gathering size restrictions in 2020.**

| state | Date issued | Date effective | gathering policy | source |
|---|---|---|---|---|
| Michigan | July 29 | July 31 | Indoor gatherings limited to 10 people | https://www.michigan.gov/whitmer/0,9309,7-387-90499_90640-535163--,00.html |
| Massachusetts | Nov 2 | Nov 6 | Indoor gatherings at private residences limited to 10, outdoor limited to 25, other limits for event venues | https://www.mass.gov/doc/covid-19-order-54/download |
| New York | Nov 11 | Nov 13 | Gatherings limited to 10 people (including private residences, indoors, and outdoors) | https://www.governor.ny.gov/news/governor-cuomo-announces-restaurants-bars-other-sla-licensed-entities-must-close-person-service |
| Minnesota | Nov 19 | Nov 20 | Most social gatherings prohibited | https://mn.gov/governor/assets/EO%2020-99%20Final%20%28003%29_tcm1055-454294.pdf |
| Illinois | Nov 18 | Nov 20 | Indoor gatherings limited to one household, outdoor to 10 people | https://coronavirus.illinois.gov/resources/executive-orders/display.executive-order-number-73.2020.html |
| Pennsylvania | Dec 10 | Dec 12 | Indoor gatherings limited to 10 people | https://www.governor.pa.gov/wp-content/uploads/2020/12/20201210-TWW-Limited-Time-Mitigation-Order.pdf |

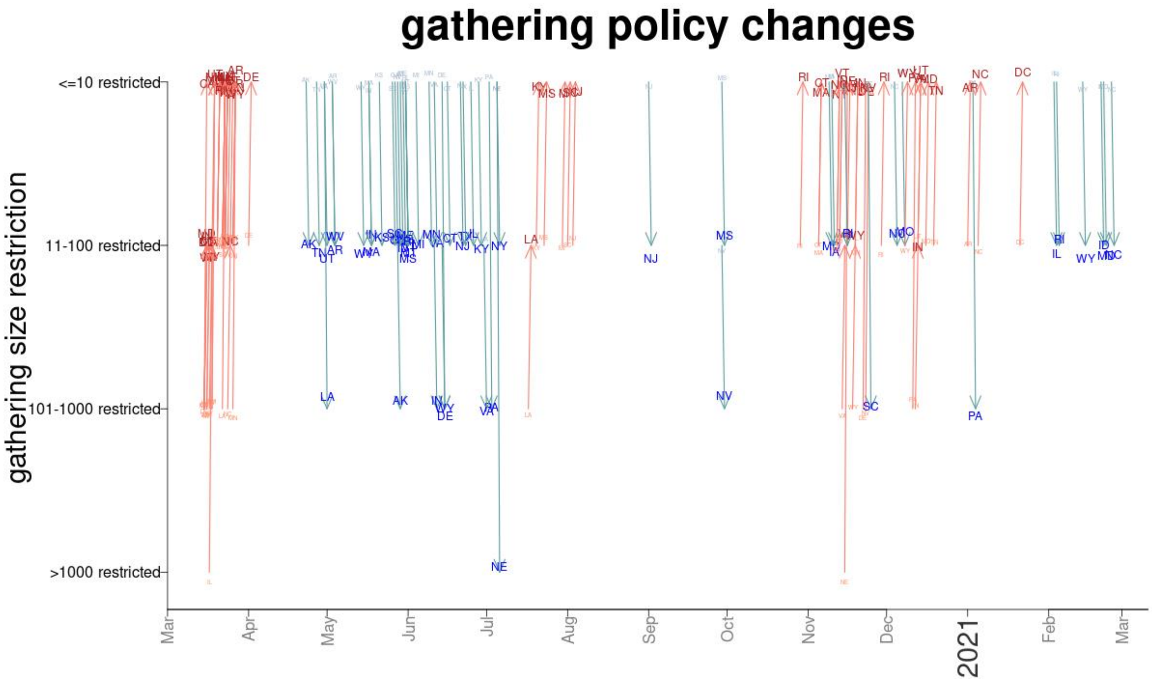


**Figure S1. Gathering size restrictions by state.** Oxford classifies statewide gathering restrictions as prohibiting gatherings >1000 people, gatherings between 101-1000 people, 11-100 people, and 1-10 people, shown on the y-axis. The red lines indicate when gathering restrictions increase, while blue is when restrictions relax, with arrows also indicating the direction. Data from the Oxford COVID-19 Government Response Tracker (https://www.bsg.ox.ac.uk/news/our-coronavirus-policy-tracker-adds-data-us-states).

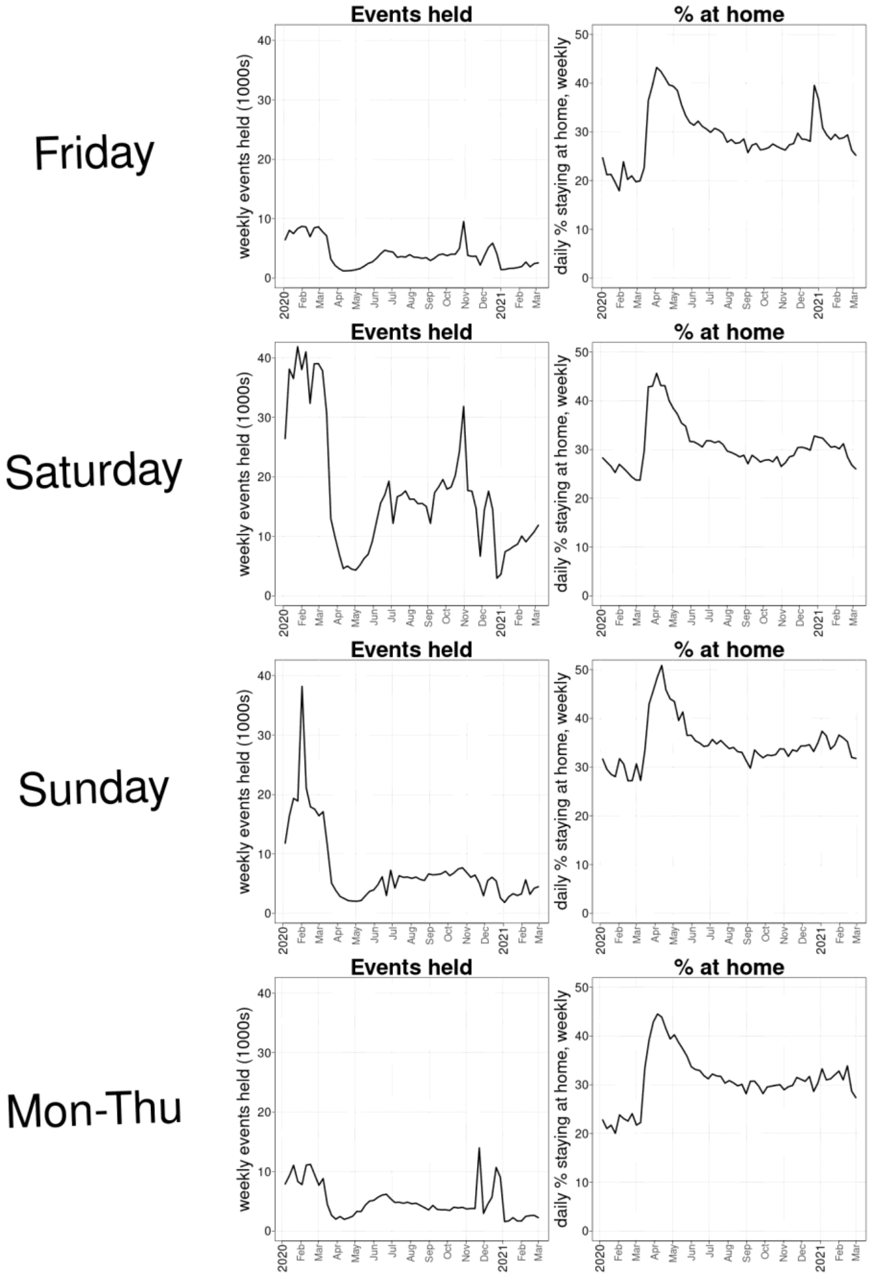


**Figure S2. Number of events held and % staying at home.** Each panel plots the weekly number of events held (left) and the weekly average % of mobile devices that stay completely at home each day (right). Each row shows a different day of the week, showing Friday, Saturday, Sunday, and Monday-Thursday data, respectively.

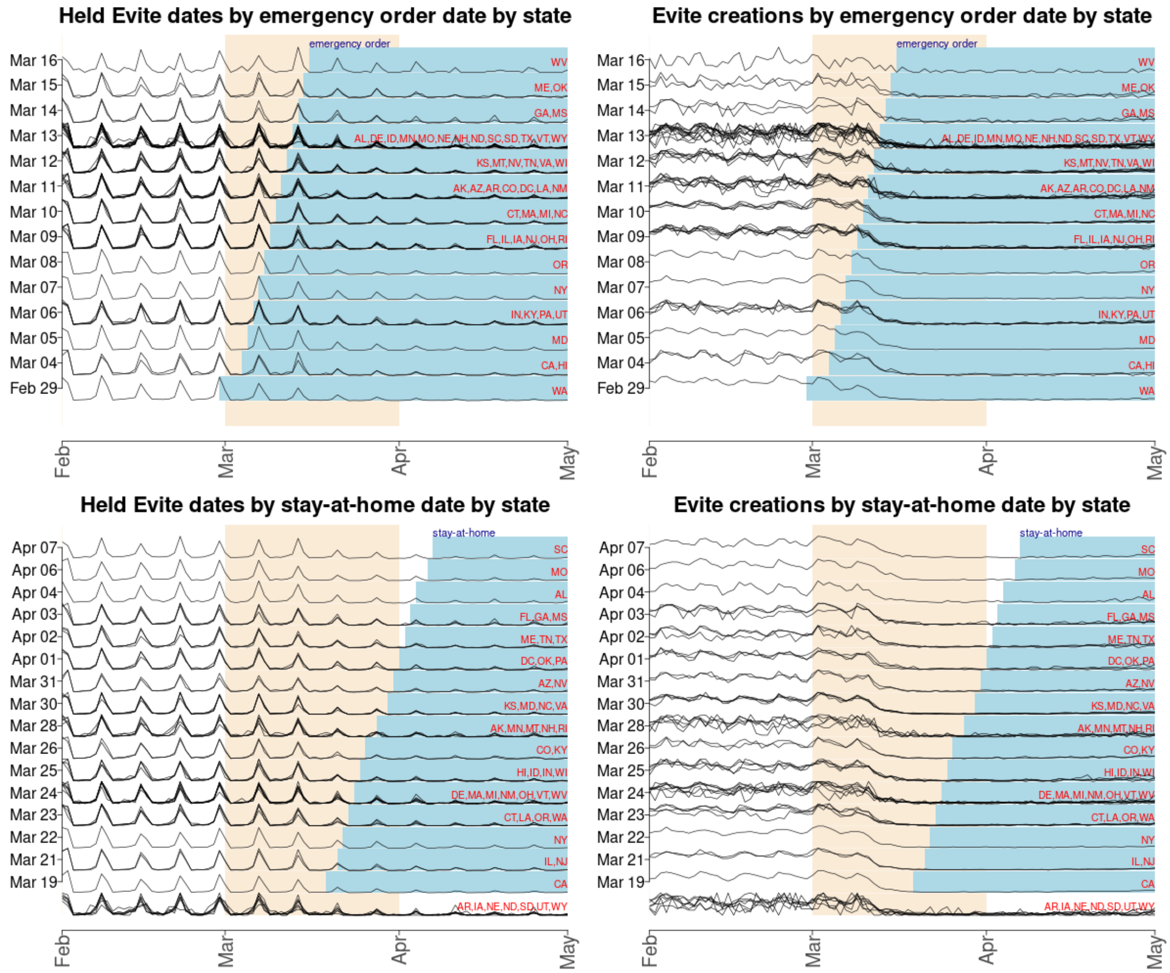


**Figure S3. Evite event and creation dates by state emergency order or stay-at-home policy date.** Top panels: states with the same emergency order date are plotted on the same row, with the state postal abbreviations red on the right. The blue bars indicate the dates where the emergency orders are in effect. The y-axes are normalized to the maximum number of events in the date range shown (February 1 through May 15 2020). Bottom panels: states with the same stay-at-home order are plotted on the same row. States that did not issue a stay-at-home order are in the bottom row. Note that the reductions in Evites that are held or planned do not correspond with the blue shaded areas at all.

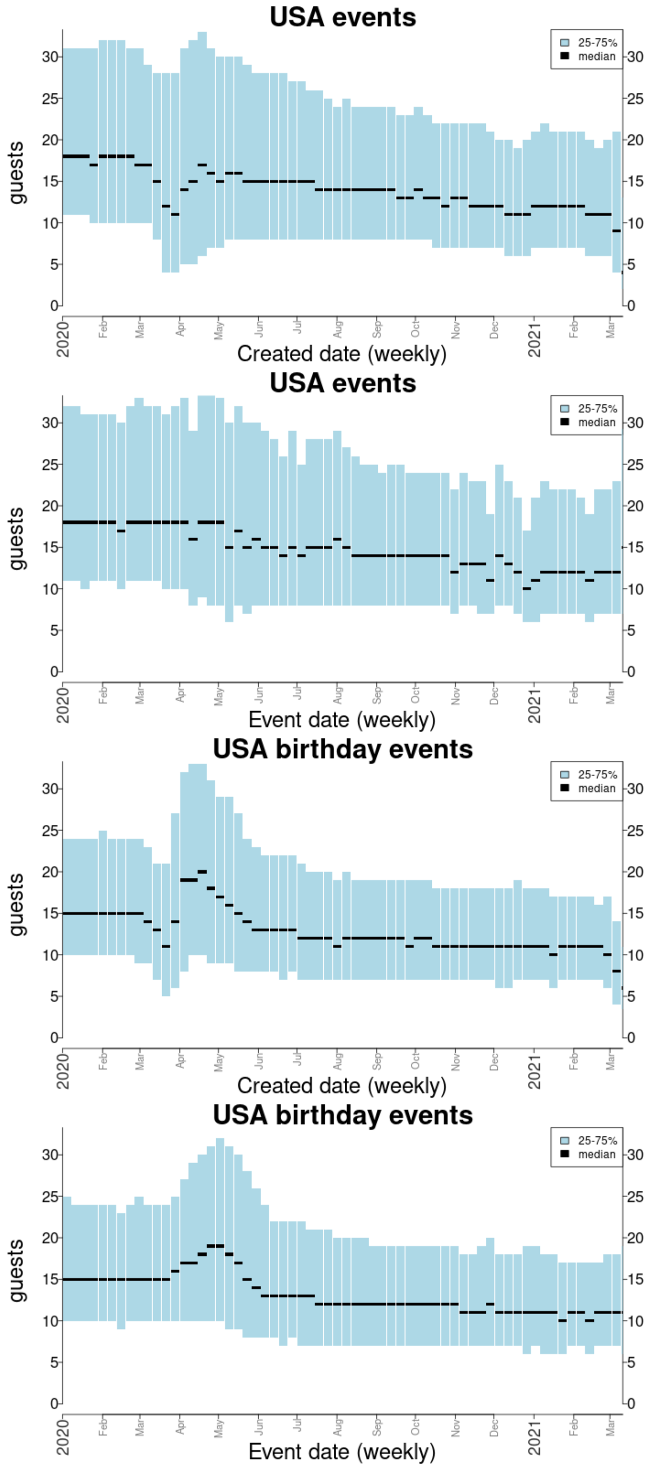


**Figure S4. Guestlist sizes over time.** Each bar captures one week, starting from Wednesday, January 1, 2020. The median event size created or organized each week is indicated in black, and the 25th to 75th percentile is indicated in blue.

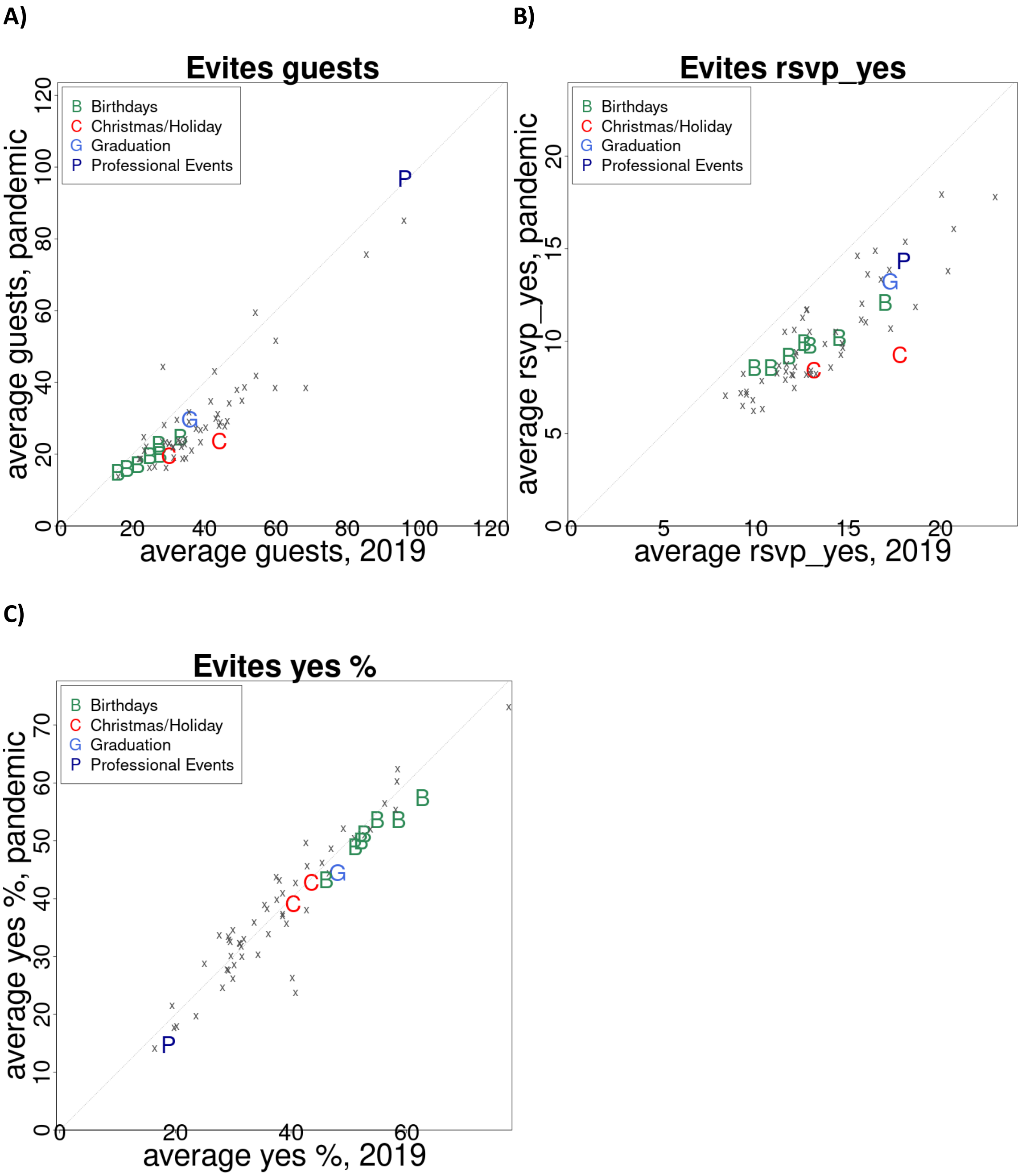


**Figure S5. Evite event sizes and RSVPs by event category in the United States.** A) Average guests invited per event by category, pandemic year vs 2019. B) Average number of guests RSVP-ing "yes" by event category, pandemic year vs 2019. C) Average % of guests RSVP-ing "yes" by event category, pandemic year vs 2019. A few event categories are highlighted, as indicated in the legends. Only categories with over 10,000 events in 2019 are plotted.

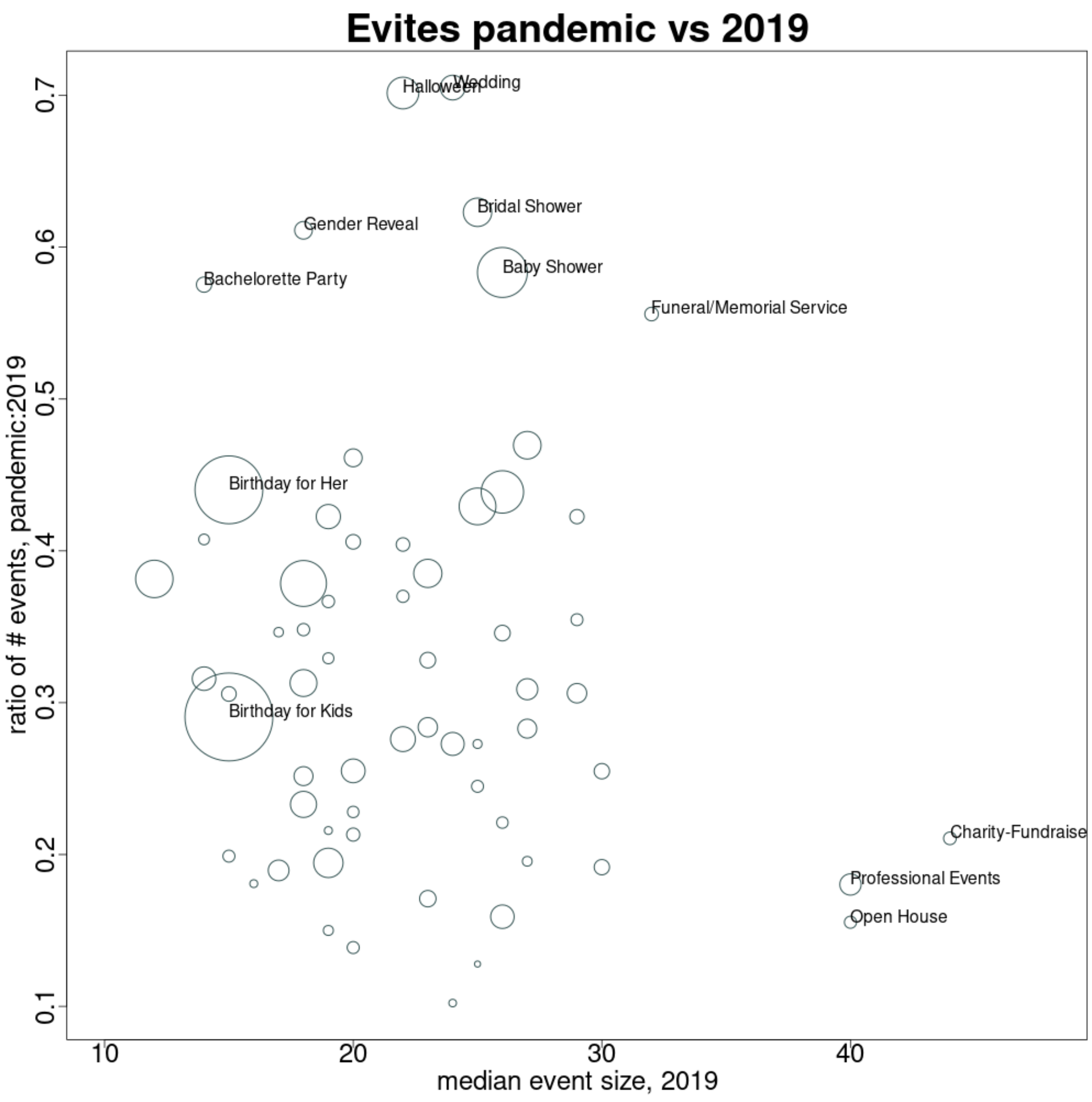


**Figure S6. Drop in number of events vs guest list size by event category.** Each dot represents an event category, with dot size proportional to the number of events during the pandemic year and selected dots labeled with the category name. The x-axis represents the median size of events in 2019 and the y-axis the ratio of events during the pandemic year vs 2019. Event categories associated with large gatherings in 2019 (professional events, fundraisers, open houses) had far fewer events during the pandemic year. Some life-change-related events, like wedding-related, births, and funerals, dropped the least. Only categories with over 10,000 events in 2019 are included in the plot.

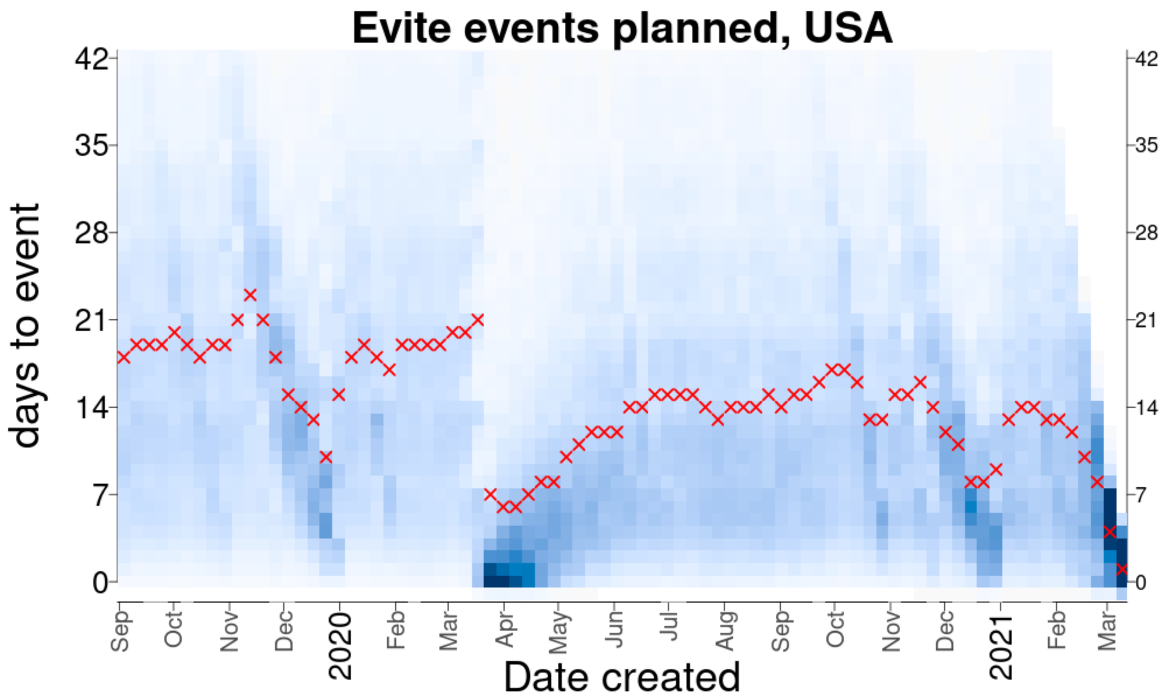


**Figure S7. Time from event creation to event date.** The red xs are the median days between Evite event creation and the scheduled event date in the US. Blue is a heatmap, with darker shades representing larger number of events planned. Results are binned at the weekly level (weeks are defined as starting on Monday). Note that before the pandemic people planned events about 2.5 weeks in advance and during the pandemic people planned events about 2 weeks in advance.

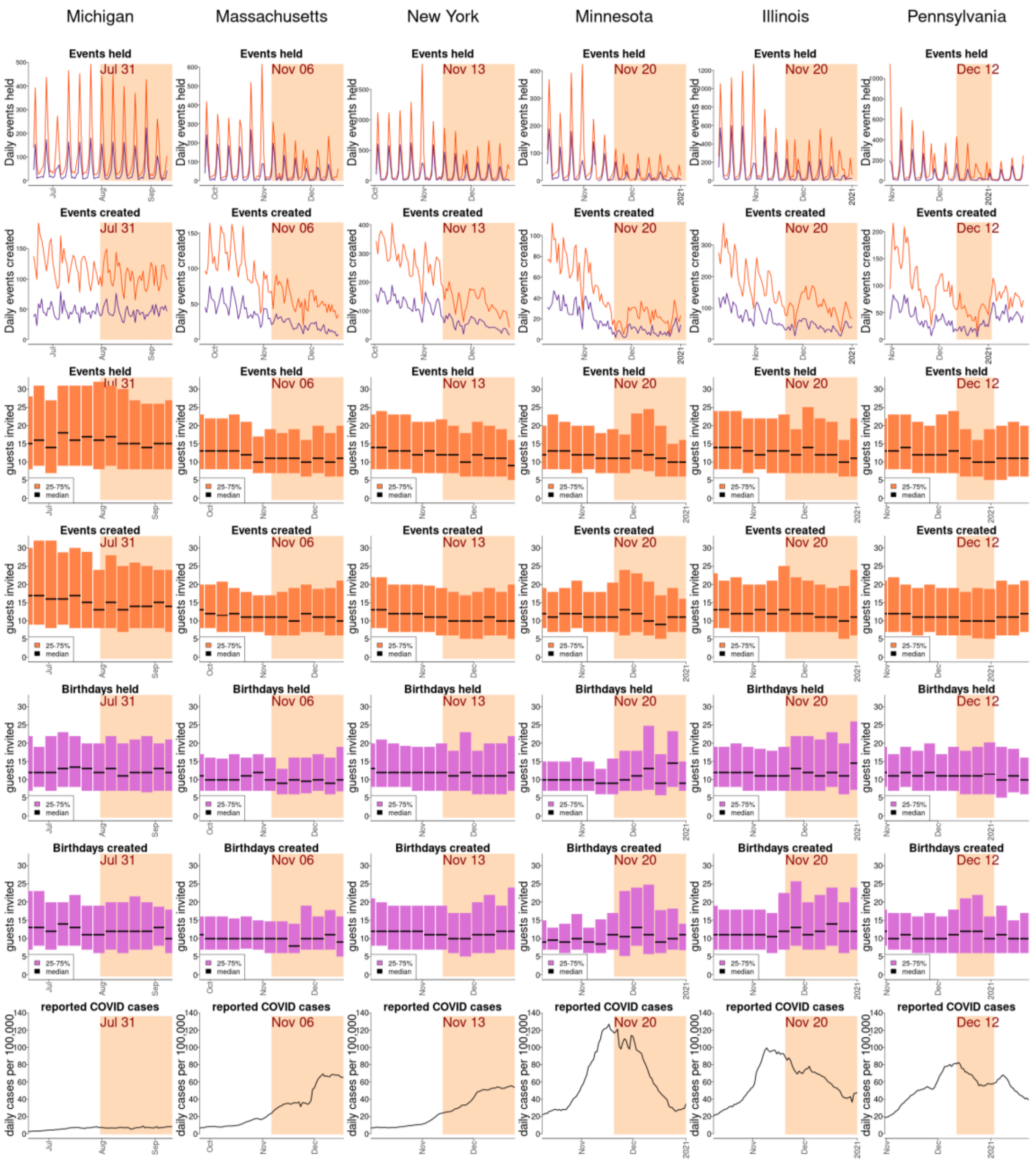


**Figure S8. Response to tightening gathering restrictions.** Each column represents a state that restricted gatherings to 10 people or fewer than 10 people. Each panel spans 12 weeks, highlighting in brown the period where gatherings >10 people were restricted. The top row is the number of events held (total events in orange and birthdays in dark purple), and the second row is the number of events created each day. The third through sixth rows are the median and interquartile interval of the number of guests invited to events. Guest numbers are summarized weekly, from Monday to Sunday. The bottom row is the smoothed number of reported COVID-19 cases per capita. Thanksgiving was on Thursday Nov 26 2020.

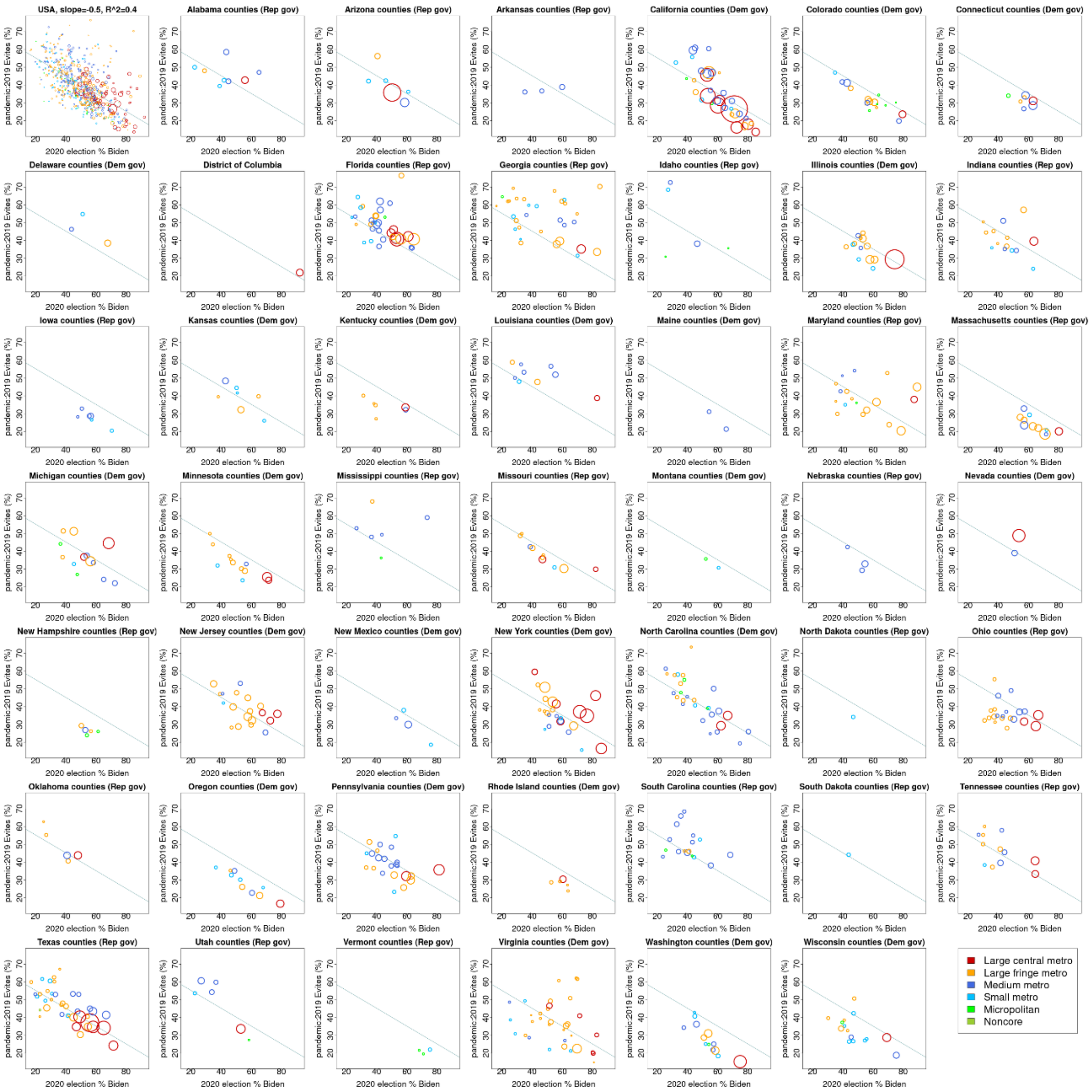


**Figure S9. Drop in Evite events during the pandemic year vs fraction voting for Biden in the 2020 election by county.** The top left panel plots the fraction of a county that voted for Biden on the x-axis vs the ratio of events during the pandemic (March 2020-2021) vs 2019. Dot sizes are proportional to county population and color to the urban/rural classification indicated in the legend. Only counties with at least 500 Evites in 2019 are plotted. The solid blue line is the linear regression fit. The other 47 panels are for individual states and the District of Columbia, but the national regression line is plotted in all panels. Party affiliation of each state's governor during 2020 is indicated in the panel titles.

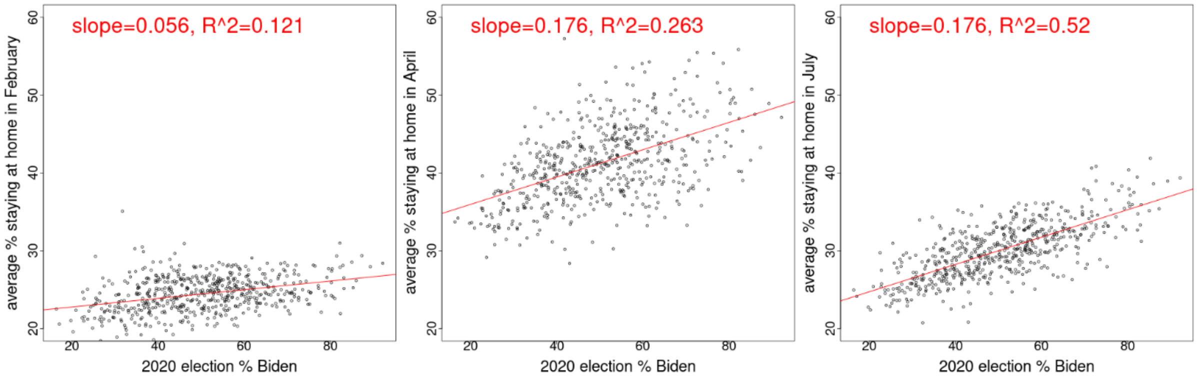


**Figure S10. Mobile devices staying at home in February, April, and July 2020 vs fraction voting for Biden in the 2020 election by county.** The average daily fraction of mobile devices staying completely at home is plotted against the fraction of each county's votes going to Biden. Only counties with at least 500 Evites in 2019 are plotted. The solid red lines are linear regression fits, and the slope and R^2 are printed in red.

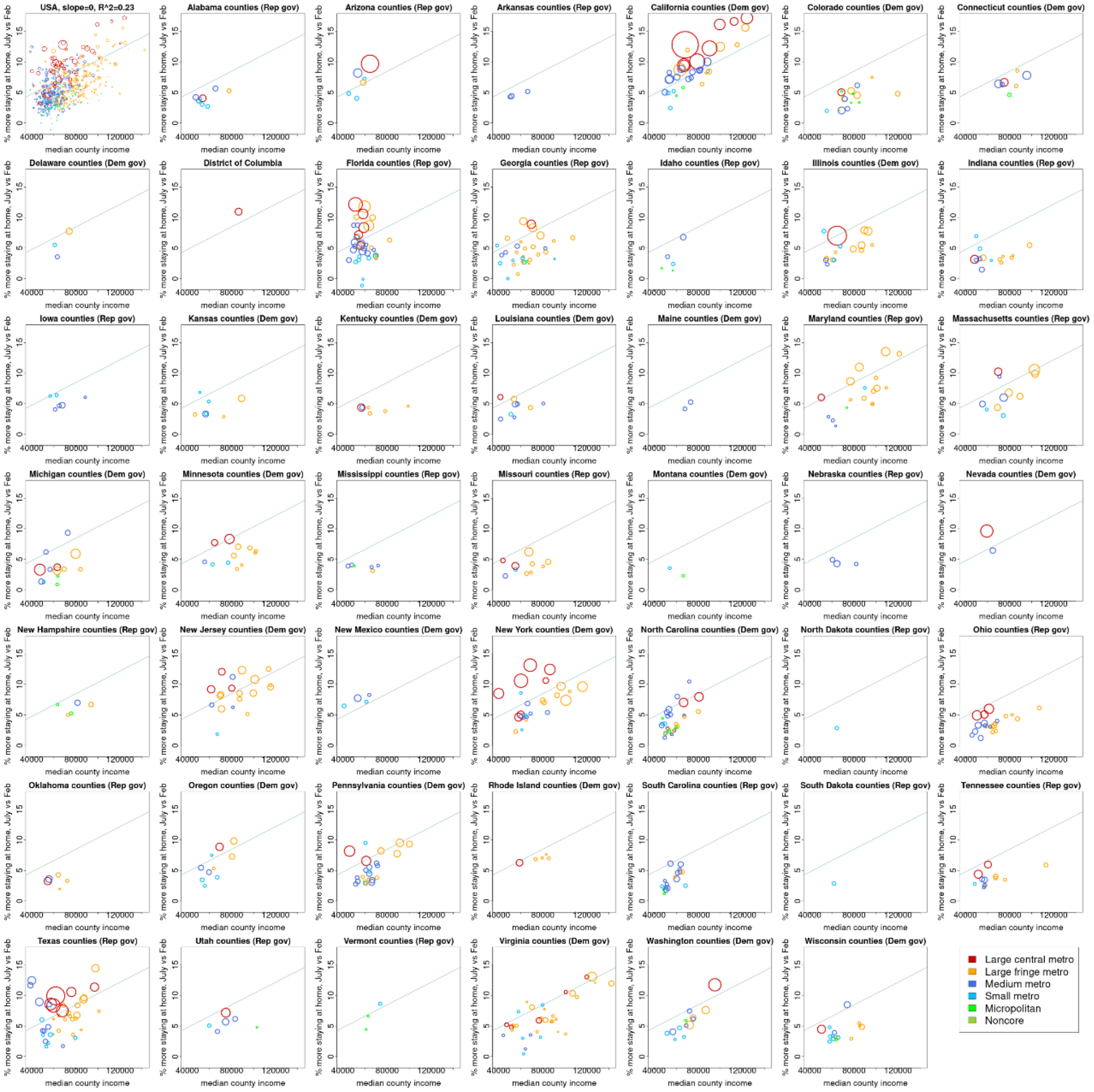


**Figure S11. Mobile devices staying at home in July 2020 vs fraction voting for Biden in the 2020 election by county.** The top left panel plots the fraction of a county that voted for Biden on the x-axis vs the average % of mobile devices staying completely at home during July 2020 compared to February 2020 (I.e., the % staying at home in July – the % in February). Dot sizes are proportional to county population and color to the urban/rural classification indicated in the legend. Only counties with at least 500 Evites in 2019 are plotted. The solid blue line is the linear regression fit. The other 51 panels are for individual states and the District of Columbia, but the national regression line is plotted. Party affiliation of each state's governor during 2020 is indicated in the panel titles.
